# ORCHID: A Comprehensive Oral Cancer Histology Image Database for Histopathological Analytics and Diagnostics

**DOI:** 10.1101/2023.08.14.23294094

**Authors:** Nisha Chaudhary, Arpita Rai, Aakash Madhav Rao, Md Imam Faizan, Jeyaseelan Augustine, Akhilanand Chaurasia, Deepika Mishra, Akhilesh Chandra, Varnit Chauhan, Rintu Kutum, Tanveer Ahmad

**Author notes:** Correspondence: Dr. Tanveer Ahmad Assistant Professor Multidisciplinary Center for Advanced Research and Studies, Jamia Millia Islamia, New Delhi-110025, India.

## Abstract

Oral cancer is a global health challenge with a difficult histopathological diagnosis. The accurate histopathological interpretation of oral cancer tissue samples remains difficult. However, early diagnosis is very challenging due to a lack of experienced pathologists and inter-observer variability in diagnosis. The application of artificial intelligence (deep learning algorithms) for oral cancer histology images is very promising for rapid diagnosis. However, it requires a quality annotated dataset to build AI models. We present ORCHID (**OR**al **C**ancer **H**istology **I**mage **D**atabase), a specialized database generated to advance research in AI-based histology image analytics of oral cancer and precancer. The ORCHID database is an extensive multicenter collection of 300,000 image patches, encapsulating various oral cancer and precancer categories, such as oral submucous fibrosis (OSMF) and oral squamous cell carcinoma (OSCC). Additionally, it also contains grade-level sub-classifications for OSCC, such as well-differentiated (WD), moderately-differentiated (MD), and poorly-differentiated (PD). Furthermore, the database seeks to bolster the creation and validation of innovative artificial intelligence-based rapid diagnostics for OSMF and OSCC, along with subtypes.

## Background and summary

Oral squamous cell carcinoma (OSCC) is a global cancer burden with a substantial number of individuals diagnosed each year, primarily in Southeast Asian countries where tobacco and associated products are commonly used^1,2^. Similarly, oral submucous fibrosis (OSMF) is a chronic and progressive condition that primarily affects the oral cavity^3,4^. It is characterized by the deposition of fibrous tissue in the submucosal layer, leading to restricted mouth opening, difficulty swallowing, and altered oral function^5^. OSMF also predominantly affects individuals in Southeast Asian countries where betel quid chewing is prevalent. The condition is known to have a potentially malignant nature, increasing the risk of developing oral cancer. Early detection and intervention are crucial in managing OSCC as well as OSMF and preventing its progression to malignancy.

The available diagnostic methods for OSMF and OSCC play a critical role in identifying and assessing these conditions. However, these methods have certain limitations that affect their accuracy and effectiveness. For OSMF, the diagnosis primarily relies on clinical examination and assessment of characteristic signs and symptoms^6^. The gold standard is a tissue biopsy and histopathology examination by a trained histopathologist. However, histopathological examination of the biopsy sample may not always provide a clear distinction between OSMF and early-stage OSCC, leading to diagnostic difficulties. In the case of OSCC, a combination of clinical examination, radiographic imaging, and biopsy is typically used for diagnosis. A clinical examination involves assessing the site, size, and appearance of the oral lesion. Radiographic imaging techniques such as computed tomography (CT) or magnetic resonance imaging (MRI) can help evaluate the extent of the tumor and identify possible metastasis^7^. Nevertheless, these imaging techniques exhibit restricted specificity when it comes to distinguishing between benign and malignant lesions.

Moreover, the lack of skilled histopathologists poses a significant obstacle, and the process of manual annotation further contributes to inter-observer discrepancies. Therefore, there is a need for further research and the development of more advanced diagnostic techniques that can improve the early detection and accurate diagnosis of these conditions, allowing for timely and appropriate management strategies to be implemented. To facilitate analysis, preprocessing of H&E images is necessary, followed by appropriate segmentation for further analysis. Computer-based algorithms have been employed to segment H&E stained images, successfully automating the process of separating the epithelial layer from the sub-epithelial layer^8^. This enables proper classification of tissue architectural changes and the extraction of relevant features for machine learning. However, despite these advancements, the application of these tools to human tissue samples has not yielded definitive results due to a lack of comprehensive histopathology databases.

To build deep learning algorithms, we need well-annotated H&E by expert histopathologists, but for oral cancer, we don’t have enough large datasets on H&E. The lack of a publicly accessible histology image database for oral diseases presents a formidable obstacle. These databases, along with digital pathology databases, play a vital role in advancing healthcare by facilitating the development of more precise AI-based diagnostic tools. They serve as valuable resources for training and refining AI models tailored specifically for healthcare applications. Several publicly available databases have been established, housing distinct image datasets for various medical conditions, thereby aiding in the training and enhancement of AI algorithms^9,10^. However, in the realm of oral cancer, the availability of image data is noticeably limited compared to other cancer types like breast, lung, and skin cancer. Existing histology image databases primarily consist of tissue slide images related to OSCC, with none specifically including OSMF. Furthermore, there is a dearth of databases containing patch-level annotated images of OSCC. While certain research groups offer low-magnification image databases, these images fail to capture intricate nuclear features, making them unsuitable for training machine learning algorithms.

While whole slide imaging (WSI) offers advantages in generating large amounts of data and capturing comprehensive tissue information, challenges such as high computational requirements, software restrictions, and costs hinder its widespread use^11,12^. Further, issues related to image quality and uniformity in WSI datasets further complicate the integration of AI-powered algorithms effectively. Moreover, the lack of publicly accessible histology image databases specifically dedicated to oral diseases poses a significant challenge^13^. There is also a conspicuous absence of high-magnification images that comprehensively represent other oral diseases. Notably, oral conditions like OSMF lack adequate representation in these databases.

Addressing this gap necessitates that we present the ORCHID database for oral cancer, with specific emphasis on conditions like OSMF and OSCC. We believe the ORCHID database will aid the scientific community in building and harnessing AI technologies to enhance the accuracy and effectiveness of AI-based diagnostic tools, ultimately improving patient care and outcomes in the field of oral healthcare.

## Methods

### Human ethical clearance

Tissue slides were collected with the approval of an ethical committee from the participating hospitals and research institutions, (1) Jamia Millia Islamia, New Delhi, (2) Maulana Azad Institute of Dental Sciences, New Delhi; (3) Rajendra Institute of Medical Sciences, Jharkhand, (4) Banaras Hindu University, Banaras, and (5) All India Institute of Medical Sciences, New Delhi, India. The buccal mucosa tissue samples were collected for three classes, normal, OSMF, and OSCC, with grade-wise annotation from the pathologists at each hospital. Data collection for the study was conducted with the explicit consent of the patients involved, following a rigorous ethical review and approval process carried out by relevant committees. Informed consent was obtained from all participants, ensuring they were fully aware of the study’s purpose, procedures, potential risks, and benefits. They were given the opportunity to ask questions and seek clarification before providing their consent to participate. Participants willingly agreed to the open publication of their data, understanding that their identities would be protected and their information anonymized. The manuscript includes specific references to ethical approval granted by different institutions, indicating their compliance with ethical guidelines and regulations. These references serve as a means of tracking and verifying the study’s adherence to ethical standards. Here are some example proposal numbers from various institutions: (Proposal No.: 6(25/7/241/JMI/IEC/2021; Proposal No:. ECR/769/INST/JH/2015/RR-18/236; Proposal No.: F./18/81/MAIDS/Ethical Committee/2016/8099; Proposal No.: IEC-828/03.12.2021). These numbers uniquely identify the respective ethical approvals received.

### Haematoxylin and eosin staining (H&E)

Biopsy samples of normal, OSMF and OSCC tissues underwent H&E staining. The staining procedure was conducted either in-house or outsourced to different laboratories. To eliminate staining variations across different laboratories, the preparation of H&E slides involved five histopathology labs, each utilizing their own independently developed and optimized protocols for the staining process. Following staining, the samples were examined under a microscope by a skilled histopathologist to assess cellular morphology, and tissue architecture, and identify any distinctive features or abnormalities specific to each sample type. This evaluation by the histopathologist involved grading the tissue slides for OSCC and OSMF, as well as differentiating between normal and diseased tissue sections. Subsequently, the annotated and validated images were utilized for further analysis.

### Image acquisition

Images were acquired using a 100X objective lens from Nikon and Leedz microimaging (LMI) bright field microscopy. Images were collected with the microscope’s NIS elements and ToupView image software, respectively. During the capture of the images from each tissue section slide, white balance adjustments were made and the camera was adjusted. We collected approximately 100- 150 images per tissue slide, which were stored in PNG file format.

### Expert annotation and validation

The data included in the ORCHID database underwent rigorous expert annotation and validation to ensure a high level of quality and accuracy. This assessment involved examining the clarity and detail of each image, ensuring that they were of a standard that allowed for accurate diagnosis and study. Images that were blurry or lacked sufficient detail were dismissed as they would not provide accurate or reliable information. Next, the experts evaluated the annotations that accompany the images. These annotations were scrutinized for consistency and accuracy, to ensure that they accurately represented the disease conditions depicted in the images. The process of labeling the slides was conducted manually by trained pathology experts. This involved a careful review of each slide to identify and label the specific disease conditions present. This procedure was crucial to ensure that the slides were correctly categorized. Furthermore, the slides that showed staining artifacts were also rejected. Staining artifacts can occur during the preparation of the slides and can alter the appearance of the tissue, potentially leading to misinterpretation or incorrect diagnosis. As such, only slides that were free from such errors and provided a clear and accurate representation of the oral pathology were included in the database.

### Stain normalization

The handling of the samples at each hospital during the collection of the tissue samples led to staining problems that persisted even after following the established H&E staining protocol. To address and minimize the variations in staining appearance across different sites in the H&E images, a stain normalization method was implemented, specifically the Reinhard stain normalization technique^14^, as shown in **Fig. 1c**. This approach, described in the study, involves a series of steps to standardize the color properties of the images to a desired standard. The first step is scaling the input image to match the target image statistics. This involves adjusting the intensity values of the input image to align with the desired color distribution of the target image. The scaling ensures that the overall brightness and contrast of the input image are consistent with the target image. The next step involves transforming the image from the RGB color space to the LAB color space proposed by Ruderman. The LAB color space separates the image into three channels: L (lightness), A (green-red color component), and B (blue-yellow color component). By performing the transformation, the image is represented in a color space that better captures the perceptual differences in human vision. Finally, Reinhard color normalization is applied to the LAB image. Reinhard color normalization adjusts the color properties of the image to align with a desired standard. It achieves this by equalizing the mean and standard deviation of the LAB channels across the image.

**Fig. 1:**
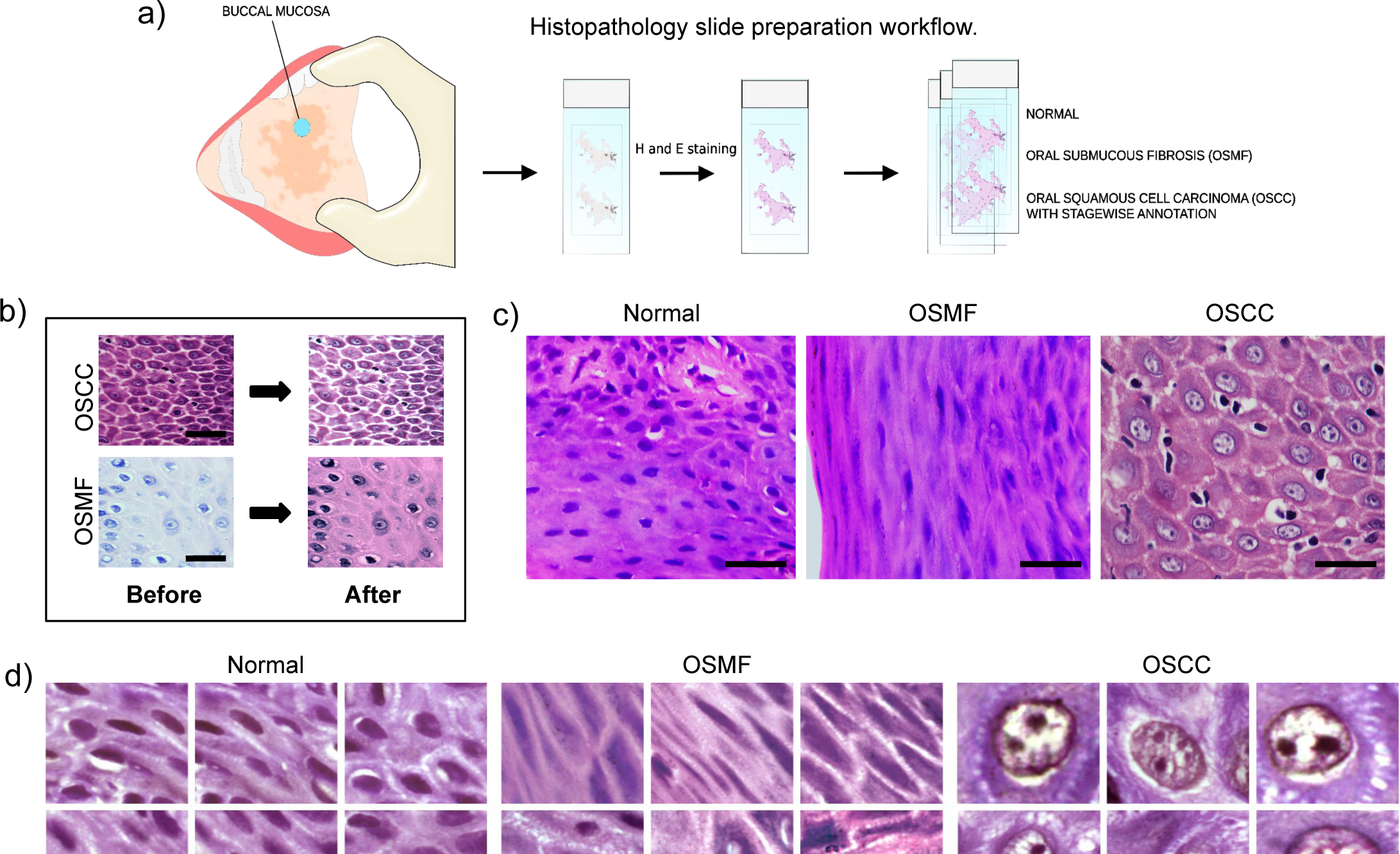
Workflow, Image Analysis, and Stain Normalization. a) The workflow for preparing oral histopathology slides involves a series of steps, from the collection of tissue samples to slide preparation and staining. b) Representative images captured at a magnification of 100X exhibit normal tissue, cases of OSMF, and cases of OSCC. These images were digitized using bright field microscopy, providing a visual depiction of the different stages involved in the preparation and staining of tissue slides. The scale bar is 10μm. c) Stain normalization is performed to standardize the stain appearance in the images. The Reinhard stain normalization method is utilized for this purpose, ensuring consistent and comparable staining across the images. The scale bar is 10μm. d) Image patches, measuring 300 by 300, are generated from the 100X images of normal tissue, OSMF cases, and OSCC cases. These patches serve as representative examples of specific regions within the larger images, offering focused insights into the characteristics of normal tissue as well as OSMF and OSCC conditions.

If the LAB statistics for the input image are not provided, they are derived from the input image itself. This ensures that the normalization process is tailored to each individual image. Below is the equation for the same:

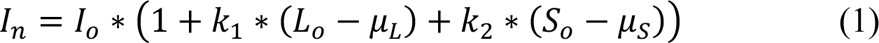

where:

I_n_ is the normalized image

I_o_ is the original image

k_1_ and k_2_ are constants that are chosen to optimize the appearance of the normalized image

L_o_ is the average brightness of the original image

S_o_ is the average saturation of the original image

μ_L_ and μ_S_ are the average brightness and saturation of a reference image.

### Patch generation

After normalization, we generated image patches of size 300 by 300 pixels from 100X objective images. The patches were generated by left-to-right sequential cropping (overlapping 150 pixels) in the original images. At this point, we also discarded those patches that have more white space, blurriness, and air bubbles reflecting image patches. The selected image patches were used as input data to build deep-learning models.

### Baseline model development and fine-tuning

We performed benchmarking of ten deep-learning algorithms through pre-training and fine-tuning our models as shown in **Fig. 2** (transfer learning). Because it had the highest accuracy among the ten pre-trained models, Inception V3 was chosen^15^. The InceptionV3 model was pre-trained on the ImageNet dataset, providing a strong initial set of learned features. The model’s top layers were excluded to allow for customization. The model was then fine-tuned by setting all layers in the Inception V3 model as trainable. This process allows the model to adapt to the specific dataset being used in the study. A flattened layer was added to convert the output of the InceptionV3 model into a 1-dimensional tensor. This was followed by a dense layer with 1024 units and a ReLU activation function, facilitating feature extraction and non-linear transformations. To mitigate overfitting, a dropout layer with a coefficient of 0.2 was introduced. Finally, a dense layer with 3 units and a softmax activation function was employed to produce the output probabilities for the three classes in the classification task. The model was compiled using the RMSprop optimizer with a learning rate of 0.0000001 (or 10e-7) and trained with the categorical cross-entropy loss function. The model’s performance was evaluated based on accuracy. For the specific task at hand, this model design combines transfer learning from Inception V3 with fine-tuning to produce an efficient image classification model. The same settings were used for both the classification models that are; the first model which classifies the image patched into normal, OSMF, and OSCC, and the second model which classifies the image patches into WD, MD, and PD grades of OSCC.

**Fig. 2:**
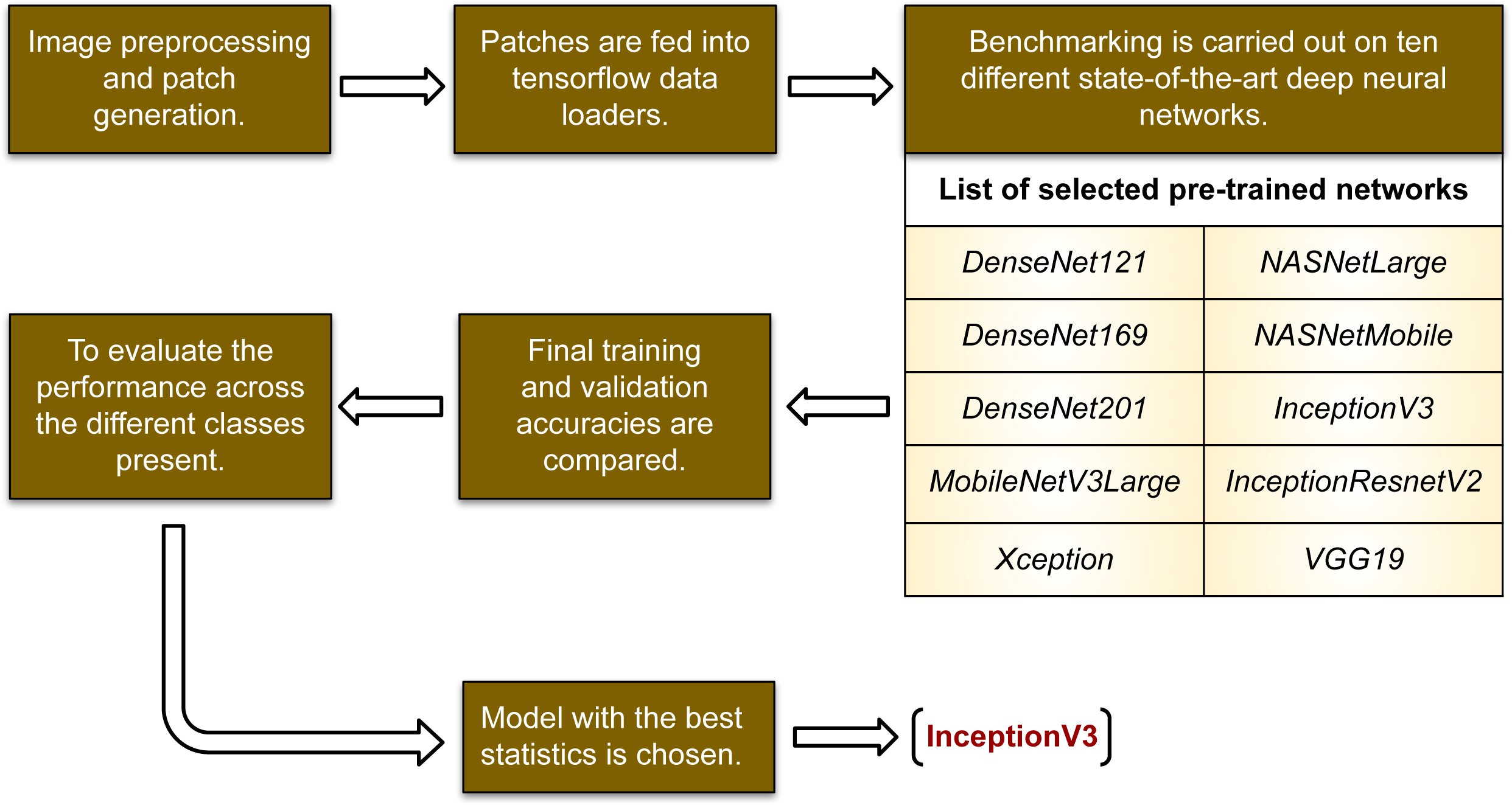
Flowchart describing the process to benchmark which pre-trained model to choose: The flowchart serves as a visual representation of the process involved in selecting an appropriate pre-trained model for a specific task. It outlines the steps and criteria to consider when evaluating different models. Further, the flowchart provides a systematic approach to benchmarking various pre-trained models, taking into account factors such as model architecture, training data, performance metrics, and compatibility with the classification task at hand.

### Data Records

The ORCHID data used in the current study has been uploaded privately through the scientific data figshare platform.

The data consists of digitized slides that were collected and stained for analysis. The digitization process involved capturing images using a 100X objective lens, as depicted in **Fig. 1a**. The dataset encompasses images from three distinct classes: normal, OSMF, and OSCC. Each class folder within the dataset contains image tiles generated at a 100X magnification level. The visual representation of these images is depicted in **Fig. 1b, 1c, and 1d,** which showcase the appearance and characteristics of the different classes. To provide a quantitative overview of the ORCHID training dataset (T.R.S.), **Table 1** presents a summary of the number of images available in each of the five classes(folders), which are as follows, Normal-TRS-TR, OSMF-TRS-TR, WDOSCC- TRS-TR, MDOSCC-TRS-TR, and PDOSCC-TRS-TR. Each class folder consists of subfolders representing different tissue slides collected from different patients. The naming of these folders start from the dataset name, ‘ORCHID’, followed by institute ID from where the sample has been collected and lastly the sample ID itself. All the image patches are stored inside these subfolders as per the tissue slide, they belong to. The naming of image patches is done in such a way, that each label represents, first the dataset name-‘ORCHID’, followed by institute ID, then sample ID, then image-ID(I) and lastly the patch-ID(P). The tabulated information helps to understand the distribution and proportion of images within each class, aiding in the analysis and utilization of the ORCHID dataset for the study or related research endeavors.

These results summarize the ORCHID database we have created, which presents a significant advancement in the field of digital histopathology for oral cancer and pre-malignant oral conditions, with a specific focus on OSCC and pre-malignant oral conditions like OSMF. Our primary objective was to develop a comprehensive high-resolution image database encompassing these conditions as well as normal healthy oral tissue. The availability of such diverse datasets is crucial for precisely classifying and differentiating between normal and diseased oral tissue. Additionally, these models will act as a standard for categorizing this dataset, and by making this information available, many researchers’ efforts to develop machine learning models for disease detection will be aided.

In summary, we have made an initial attempt to provide a comprehensive image database for two of the most prominent oral conditions, OSCC and OSMF. We believe that more such databases will be made publicly available in the near future. These comprehensive image databases will facilitate the development of accurate AI-based diagnostic tools for oral diseases, ultimately improving patient care and outcomes in the field of oral healthcare. In future, integration of databases comprising molecular markers, transcriptome, metabolome, and other biomarkers, combined with oral histological image through advanced AI-driven imaging techniques, holds great promise in improving diagnostic accuracy and precision. This potential has already been observed in the diagnosis of lung and breast cancers.^16^.

### Technical Validation

The histology images in the ORCHID database involved a rigorous and systematic approach to ensure the reliability and accuracy of the dataset. To validate the dataset, a subset of images was randomly selected, which was then used as input for convolutional neural network (CNN) algorithms. Specifically, the chosen architecture for the CNN models was Inception V3, and the details of its configuration and implementation can be found in the methods section. The selection of the Inception V3 model was based on a comparison of training and validation accuracies among ten state-of-the-art neural network models. It was found that the Inception V3 model performed the best among all the models, achieving the highest accuracy in classification tasks (**Fig 4)**. The results demonstrated that the model successfully distinguished between the normal, OSMF, and OSCC classes, with a training accuracy of 98.54% and a testing accuracy of 97.15%, as illustrated in **Fig. 3a**. However, when classifying the three different grades of OSCC (WD, MD, and PD), the classification accuracy was slightly lower. The model achieved a training accuracy of 87.54% and an internal validation accuracy of 61.42%, as shown in **Fig. 3b**. These results indicate that the model had relatively higher difficulty accurately classifying the different grades of OSCC compared to the overall classification task. The technical validation process ensures that the ORCHID database is reliable and suitable for subsequent analysis and research. The performance of the Inception V3 model on the dataset demonstrates its capability to accurately classify normal, OSMF, and OSCC cases. Nevertheless, the performance in classifying the different grades of OSCC indicates the potential for improvement and calls for additional investigation and refinement. Furthermore, the need for a substantially larger image dataset is recognized, and efforts toward expanding the dataset are currently underway as part of ongoing work.

**Fig. 3:**
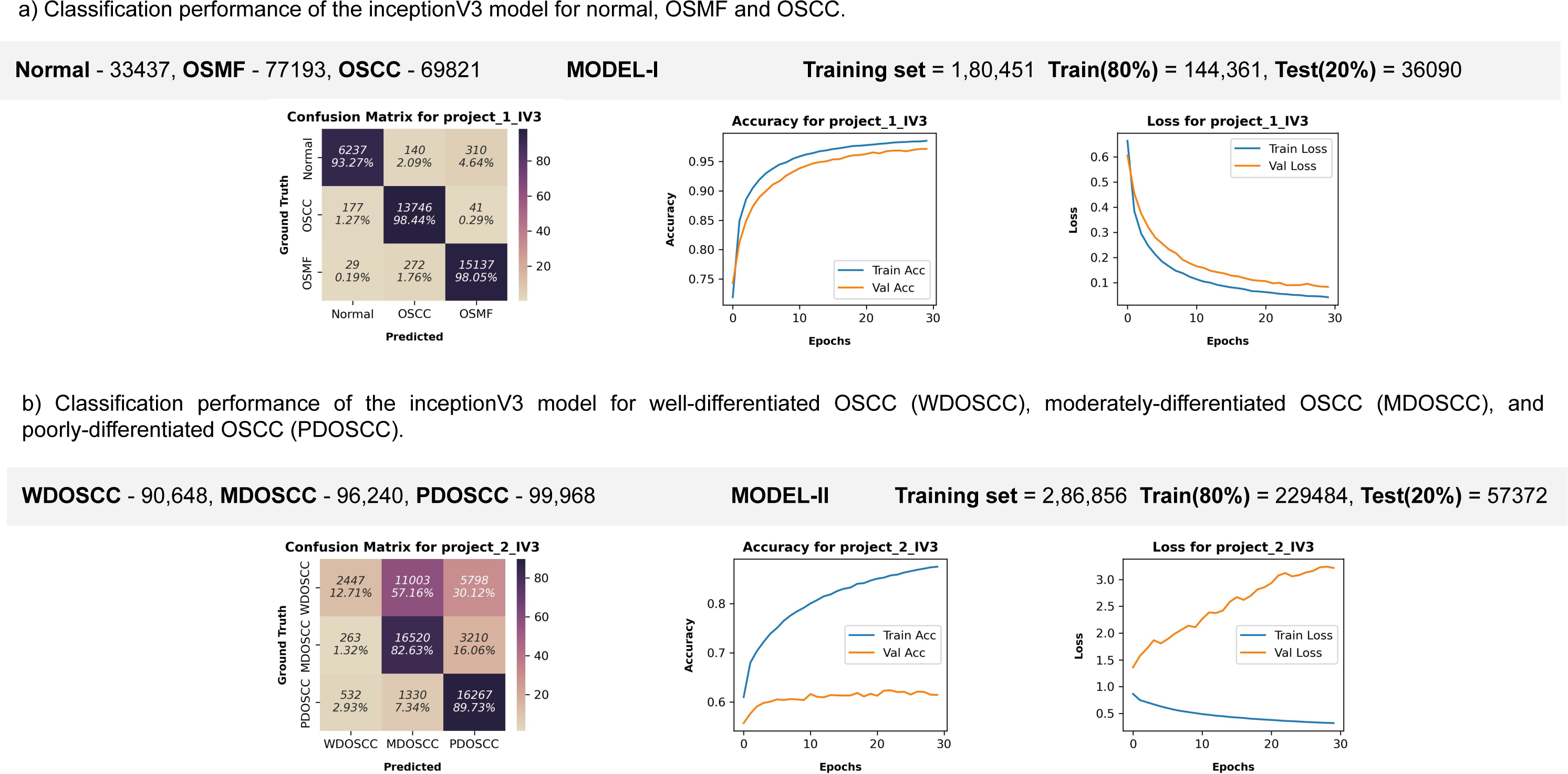
Models performance evaluation metrics. a) **Classification performance of the inceptionV3 model for normal, OSMF and OSCC.** This model focus on assessing the effectiveness of classification algorithm Inception V3 (IV3) in distinguishing between different oral tissue conditions: Normal, OSMF, and OSCC. The metrics provide a quantitative measurement of the accuracy, prediction performance (confusion matrix) and loss for value representing the training loss and validation loss. b) **Classification performance of the inceptionV3 model for well-differentiated OSCC (WDOSCC), moderately-differentiated OSCC (MDOSCC), and poorly-differentiated OSCC (PDOSCC).** This model specifically focus on the classification of different grades of OSCC: well-differentiated OSCC (WDOSCC), moderately-differentiated OSCC (MDOSCC), and poorly-differentiated OSCC (PDOSCC). The metrics measure the performance of classification algorithm, Inception V3 (IV3) in correctly classifying and differentiating between these different grades of OSCC.

**Figure 4:**
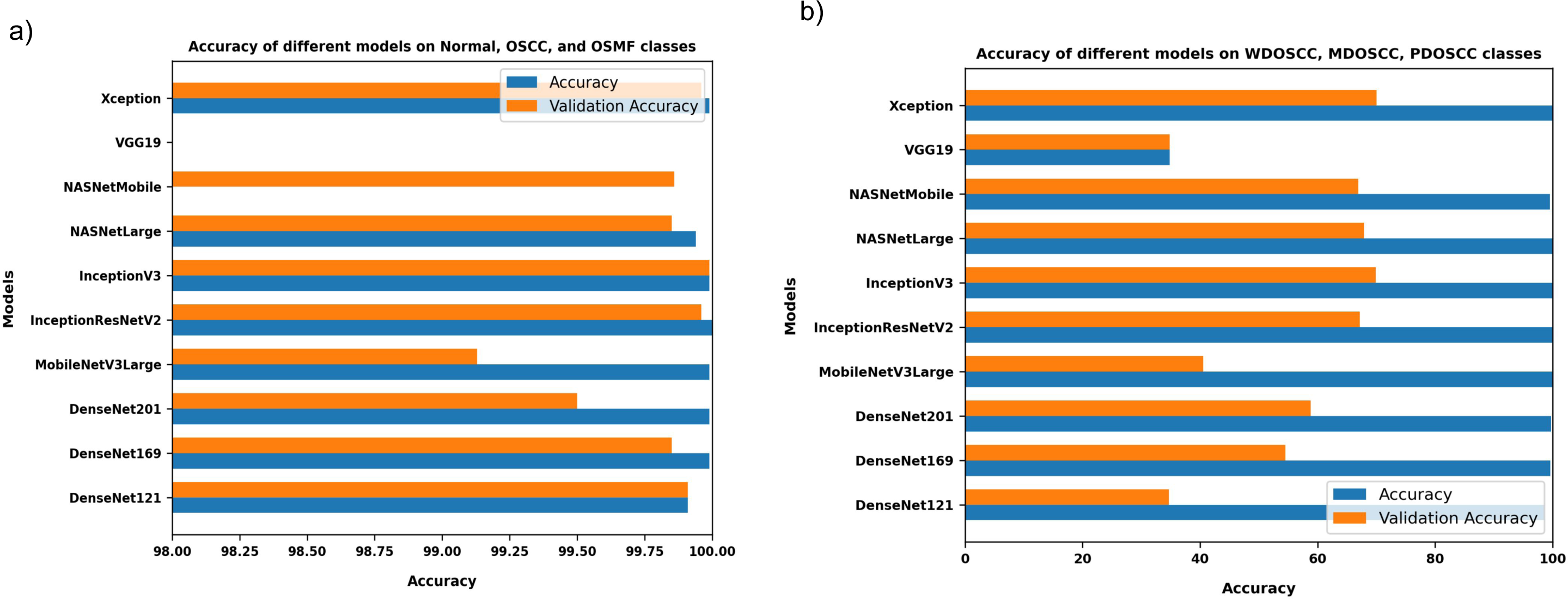
Histogram showing accuracy comparison of ten pre-trained models benchmarked for the study. a) Accuracy of the selected ten pre-trained models on Normal, OSMF and OSCC image patches. b) Accuracy of the selected ten pre-trained models on well-differentiated OSCC (WDOSCC), moderately-differentiated OSCC (MDOSCC), and poorly-differentiated OSCC (PDOSCC).

## Data Availability

All data produced in the present study are available upon reasonable request to the authors

## Acknowledgements

N.C. is the recipient of a senior research fellowship from the Indian Council of Medical Research(3/1/2(1)/Oral/2021-NCD-II), New Delhi, India. This work was also supported by the Science and Engineering Research Board (CRG/2020/002294), and the Indian Council of Medical Research (ICMR) (GIA/2019/000274/PRCGIA (Ver-1)), New Delhi, India. We also acknowledge the computing support from the Mphasis F1 Foundation and the Center for Bioinformatics and Computational Biology (B.I.C.) (BT/PR40220/BTIS/137/22/2021) facility at Ashoka University. We are thanking Farhat Zeba and Sumra Khan for helping out with the imaging.

## Author contributions

N.C. and T.A. conceptualized the study. A.R. and M.I.F. collected and processed tissue slides. N.C., M.I.F., and V.C. performed microscopy imaging. N.C. and A.M.R. performed image processing, CNN training, designed the ORCHID, and analyzed the results. A.R., J.A., AA.C., D.M., and A.C. annotated the oral cases and provided their pathology expertise and guidance. R.K. and T.A. supervised the study. All the co-authors contributed feedback and suggestions towards the preparation of the manuscript.

